# International patient safety learning systems: a qualitative key informant interview study with safety-critical industry experts

**DOI:** 10.64898/2026.07.27.26359014

**Authors:** Jaafer Qasem, Fiona Wood, Adrian Edwards, Andrew Carson-Stevens

## Abstract

**Background:** Patient safety learning systems operate primarily at the national level, yet the potential for international sharing of safety data and learning remains largely unrealised. Other safety-critical industries, including civil aviation, nuclear power, and rail, have developed international incident reporting and learning systems whose design principles may offer transferable insights for healthcare.

**Objective:** To explore the views and experiences of safety-critical industry experts regarding the purpose, key functions and features, transferability, barriers, enablers, and incident priorities of a potential international patient safety learning system.

**Design:** Qualitative phenomenological study using semi-structured key informant interviews analysed with the five-stage framework analysis method.

**Participants:** Eleven international experts purposively sampled from safety-critical industries (healthcare, civil aviation, maritime, and railway) across six countries. Interviews were conducted by telephone or internet-based audio call between May and September 2019.

**Results:** Seven themes were identified: (1) purpose of an international patient safety learning system; (2) key functions; (3) key features; (4) transferability of learning; (5) enablers; (6) barriers; and (7) patient safety incident types appropriate for international sharing. Rare and emerging event detection emerged as the most distinctive value of an international system, a capability that cannot be replicated at national level. Critical functions included robust feedback loops, multidisciplinary analytical capability, and actionable recommendations. Key barriers, including blame culture, inter-organisational mistrust, and cross-border privacy legislation, were each amplified at the international level relative to their national-level equivalents.

**Conclusions:** This study provides the first cross-industry qualitative account of design requirements for an international healthcare patient safety learning system, highlighting transferable lessons from established industries alongside healthcare-specific challenges that require bespoke solutions.

## Introduction

Patient safety incidents result in preventable harm to millions of patients worldwide each year. The Canadian Adverse Events Study estimated that approximately 7.5% of hospital admissions involved adverse events, and comparable figures have been reported internationally (Baker et al., 2004; Kohn et al., 2000; World Health Organization [WHO], 2017). National patient safety learning systems have been established in many countries to capture incidents and generate learning, yet they operate largely in isolation: each country analyses its own incidents against its own denominators, limiting the ability to detect rare safety signals that only become visible at scale and to disseminate learning rapidly across borders.

The potential of international safety data sharing is well established in other safety-critical industries. The International Civil Aviation Organization has mandated a global safety data collection and management framework since the 1990s, enabling cross-national detection of rare events and dissemination of safety recommendations across 193 member states (ICAO, 2016). In healthcare, no equivalent international structure exists. The WHO Global Patient Safety Action Plan 2021–2030 calls for stronger international safety data sharing (WHO, 2021), yet fundamental questions remain about the purpose, functions, features, and implementation of such a system.

Existing literature on national patient safety learning systems identifies feedback mechanisms, anonymity, and ease of reporting as critical design features (Health Quality Ontario, 2017; Klemp et al., 2015; Stavropoulou et al., 2015). However, comparable qualitative evidence at the international level, drawing on practitioners from other safety-critical industries, does not exist. A prior systematic literature review, conducted as the first phase of the broader doctoral research programme that generated this study, confirmed this gap (Qasem et al., in preparation). Semi-structured key informant interviews with internationally recognised experts were therefore conducted, aiming to develop an in-depth understanding of the purpose, key requirements, and anticipated challenges of an international healthcare patient safety learning system.

## Methods

### Study design

A qualitative phenomenological design was adopted, consistent with the aim of generating in-depth expert perspectives on an underexplored topic (Creswell & Poth, 2016). The study constituted the second phase of a sequential exploratory mixed-methods doctoral research programme; its findings subsequently informed a modified Delphi consensus study with international healthcare experts (Qasem et al., 2026).

### Participants

Purposive sampling was used to identify internationally recognised experts with direct experience of incident reporting and learning systems in safety-critical industries. Eligibility required senior professional experience in a relevant safety-critical industry and direct involvement in the design, governance, or evaluation of incident reporting and learning systems. Experts were identified through systematic literature review reference lists, WHO consultative meeting participant lists, and snowball referrals. Of 21 experts invited, 18 replied and 11 consented and completed interviews, a response-to-completion rate of 61% (11/18); sampling continued until thematic saturation was reached (Fusch & Ness, 2015). Table 1 summarises participant characteristics.

**Table 1.** Participant characteristics.

| ID | Industry | Country | Area of expertise |
| --- | --- | --- | --- |
| P1 | Healthcare / Aviation | UK | International IRLS design; cross-industry learning |
| P2 | Maritime | Finland | Database design; international incident classification |
| P3 | Healthcare / Aviation | UK | Aviation safety; cross-industry applications |
| P4 | Healthcare | UK | Quality improvement; safety culture |
| P5 | Healthcare | UK | Patient safety learning; knowledge mobilisation |
| P6 | Healthcare | Australia | International patient safety learning system feasibility |
| P7 | Healthcare | UK | Serious adverse events; never events; NHS executive |
| P8 | Healthcare | UK | Patient safety research; international reporting policy |
| P9 | Healthcare | Canada | Patient safety policy; WHO/ICPS governance |
| P10 | Healthcare | UK | Patient and public involvement in safety |
| P11 | Healthcare | Norway | Incident classification; Nordic reporting systems |

### Data collection

Semi-structured interviews were selected to allow the researcher to follow a predetermined guide while remaining responsive to participant-led elaboration, a balance particularly valuable when interviewing elite experts (Bearman, 2019; DeJonckheere & Vaughn, 2019). The interview guide covered six topic areas: the purpose of an international patient safety learning system; key functions; key features; transferability of learning; barriers and enablers; and incident types appropriate for international sharing. Two structurally identical versions were created, one for healthcare participants and one for those from other safety-critical industries, ordered to begin from each participant’s primary industrial context. The guide was piloted and amended prior to data collection. Both versions are provided as Supplementary File 2.

All interviews were audio-recorded with participant consent and transcribed verbatim by a university-approved professional transcription service, then proofread by the lead researcher. Duration ranged from 31 to 50 minutes (mean: 35 minutes). Non-specific participant codes (P1–P11) are used throughout to fulfil participants’ request for anonymised attribution.

### Data analysis

Framework analysis was used to identify, describe, and interpret patterns across transcripts (Ritchie & Spencer, 1994). It was selected for its explicit link between data interpretation and study aims and its accommodation of both deductive and inductive coding simultaneously, an appropriate fit for the pragmatic paradigm underpinning this study. The five stages were followed sequentially: familiarisation; thematic framework development; indexing using NVivo (version 12.1.90); charting into a thematic matrix; and mapping and interpretation. An extract of the analytical matrix is provided as Supplementary File 1. To reduce the risk of subjectivity, two transcripts (approximately 20% of the dataset) were independently coded by an experienced qualitative researcher on the supervisory team; discrepancies were resolved by consensus.

### Reflexivity

Reflexive practice was integral throughout (Berger, 2015; Dodgson, 2019). The lead researcher held a medical background and a prior scholarly interest in incident reporting and learning system development, positions acknowledged as potentially shaping data generation and interpretation. The power dynamics were notably inverted relative to conventional interview research: the lead researcher was a doctoral student interviewing internationally recognised experts with far greater substantive knowledge. This was managed through a structured interview guide, studied neutrality, probing rather than directive questioning, and reflexive memos throughout data collection and analysis.

### Ethics

Ethical approval was obtained from the Cardiff University School of Medicine Research Ethics Committee (SMREC Reference Number: 18/70). All participants provided written informed consent. Data were stored on a password-protected institutional server. All quotations are anonymised.

## Results

Eleven experts participated from safety-critical industries and countries spanning six nations (Table 1). Participants had worked for national health services, national audit bodies, the WHO, Departments of Health, and international patient safety bodies, with several holding or having held board-level positions including on the WHO’s International Classification for Patient Safety Development Committee. Seven themes emerged from framework analysis; Table 2 provides an overview with sub-themes and their contribution to existing literature.

**Table 2.** Summary of themes, sub-themes, and contribution to existing literature.

| Theme | Sub-themes | Contribution to existing literature |
| --- | --- | --- |
| 1. Purpose | Facilitating learning; awareness; improving existing systems; error detection; standardisation | Rare and emerging event detection as the distinctive international value-add: not identified in national patient safety learning system literature (Klemp et al., 2015; Stavropoulou et al., 2015) |
| 2. Key functions | Feedback mechanisms; investigative and analytical capability; generation of safety recommendations | Feedback identified as the most consistently failing function; multidisciplinary analytical teams including human factors not previously articulated for international systems |
| 3. Key features | Broad database; ease of access; universal guidelines and definitions; simplicity of reporting; anonymity; ease of classification; regular updates | Single nationally responsible organisation; open-access recommendation repository; incident-versus-solution database distinction: all new to healthcare patient safety learning system literature (Harsoor, 2010; Donaldson, 2009, 2011) |
| 4. Transferability of learning | Common taxonomy and reporting mechanisms; shared learning forums; context-sensitivity of recommendations | Standardisation–contextualisation tension as a structural design challenge: novel finding not in national literature |
| 5. Enablers | Sharing learning and developing recommendations; quality of investigation; knowledge mobilisers; legislative protection | Legislative protection for reporters drawing on aviation law: not previously advocated in healthcare patient safety learning system literature |
| 6. Barriers | Blame and organisational culture; trust and transparency; involvement and engagement; privacy legislation; data interest | Privacy legislation and culture–trust nexus amplified at international level; barriers identified as contingent rather than inherent (Weaver et al., 2013) |
| 7. Patient safety incidents | Serious adverse events and never events; rare and emerging incidents; high-risk medication incidents; near-misses with high harm potential | First empirical prioritisation of incident types for an international healthcare patient safety learning system; rare events as the most distinctive international value (Howell et al., 2017) |

### Theme 1: Purpose

All participants endorsed improving patient safety through systemic learning as the overarching purpose. Four sub-themes elaborated this purpose: facilitating cross-border awareness of incidents before they occur domestically; improving existing national systems through international benchmarking; standardising reporting terminology as a prerequisite for cross-national aggregation; and, most distinctively, detecting rare and emerging incidents. This final sub-theme had the least precedent in the existing literature. No single national system accumulates sufficient cases to detect or characterise rare events; an international system provides the case volume necessary to do so:

> *“For the international level, the main purpose is I think to detect rare and emerging incidents, the purpose of the system is to be less focused on the really big problems that we know exist, but more on rare or emerging incidents.”*

### Theme 2: Key functions

A robust, timely feedback loop was the most emphatically endorsed functional requirement. All participants regarded feedback as essential for sustaining reporter engagement, and the consensus was sharpened by direct experience of its failure in current national systems (Hewitt et al., 2017; Stavropoulou et al., 2015):

> *“Feedback mechanisms are critical in kind of local reporting systems, but they don’t work very effectively currently, and often that’s a complaint that reporters have: that they report, they spend time, they make the effort to do it and they never hear anything about it again.”*

A multidisciplinary analytical team including human factors expertise was identified as a functional requirement with no direct equivalent in the existing national patient safety learning system literature. The translation of incident analysis into clear, quality-controlled recommendations was identified as the defining output function.

### Theme 3: Key features

A broad, well-maintained database with openly accessible recommendations was the most frequently described structural feature. Participants drew a specific distinction between a database of reported incidents and a database of safety solutions, both regarded as necessary but serving different functions. All participants identified anonymity as non-negotiable, viewing it as the foundation of a blame-free reporting culture:

> *“I’d prefer anonymity. Anonymity lowers the barriers in reporting. This way all information is better handled, available and easier to share when there are not restricting confidentiality issues.”*

Ease of access was identified as a prerequisite rather than an aspiration. One participant proposed designating a single nationally responsible organisation to manage access and oversight, combining simplicity at reporter level with clear national governance, a model with no direct equivalent in current healthcare patient safety learning system literature. An agreed, high-level classification system and regular database updates were also identified as essential features.

### Theme 4: Transferability of learning

Transferability was identified as contingent on shared taxonomies, common reporting mechanisms, and structured opportunities for cross-national exchange. Despite enthusiasm for learning from aviation and nuclear models, most participants were emphatic that healthcare’s complexity precludes direct transplantation. This generated a structurally significant tension not identified in the existing literature: standardisation of reporting processes is necessary for valid cross-national comparison, while contextualisation of recommendations is necessary to avoid inappropriate generalisation to settings where the incident context differs fundamentally:

> *“Even then I think it may need to be contextualised according to different regions, so for instance if you were to have a learning and reporting system for Western Europe, the priorities of that particular area would be completely different.”*

Participants identified this tension as a distinctive design challenge for an international healthcare patient safety learning system.

### Theme 5: Enablers

Cross-border information sharing, translating incident reports into clear, quality-assured recommendations, was identified as the primary enabling condition, dependent on the credibility and contextual relevance of the outputs to potential recipients. The quality of the investigation process, not just the reporting mechanism, was a distinct enabler: an international governance body with cross-national representation was seen as the enabling structure. Legislative protection for reporters, ensuring no punitive consequences for reporting, was identified as the most fundamental systemic enabler, drawing directly on the aviation model:

> *“Legislation, if you have legislation that ensures that reporters are not punished, then that can be an enabling factor.”*

This advocacy for a legislative framework for healthcare reporting protection has no precedent in the national patient safety learning system literature.

### Theme 6: Barriers

Organisational blame culture was the most pervasive barrier, operating as a compound mechanism: blame deters reporting, absence of reporting impoverishes data, poor data quality undermines trust, and lack of trust reinforces disengagement from the system (Weaver et al., 2013). Frontline staff disengagement, driven by competing time pressures and the absence of feedback from national systems, was identified as a structural barrier amplified at the international level. One participant offered a theoretically significant observation: barriers are not fixed properties. A cultural or legislative barrier can be transformed into an enabler under the right conditions, pointing to their contingent rather than inherent nature. Cross-border privacy legislation was identified as a barrier qualitatively amplified at the international level, introducing jurisdictional complexity around data access rights that national data protection frameworks do not address.

### Theme 7: Patient safety incidents appropriate for international sharing

Experts converged on four incident categories as priorities: serious adverse events and never events, identified as the logical starting point because of their severity and tendency to recur across settings; rare and emerging incidents, identified as the category most distinctively served by the international level because of the case volume required for detection; high-risk medication incidents; and near-misses with high harm potential, selected by harm potential rather than actual harm:

> *“Well, I don’t think it needs to have resulted in harm, but it just has to have the potential for it.”*

This constitutes the first empirical prioritisation of incident types for an international healthcare patient safety learning system.

## Discussion

### Statement of principal findings

This study provides the first cross-industry qualitative account of design requirements for an international healthcare patient safety learning system. Seven themes emerged from framework analysis with expert practitioners from healthcare, civil aviation, maritime, and railway sectors across six countries. Rare and emerging event detection is the primary distinctive value of an international system: no national system can replicate this capability because the case volume required to detect and characterise rare events is only achievable at scale. Feedback loops, multidisciplinary analytical capability including human factors expertise, and actionable recommendations are the core functional requirements. Blame culture, cross-border privacy legislation, and inter-organisational mistrust are barriers amplified at the international level. Legislative protection for reporters, drawing on the aviation model, emerges as a novel enabling condition not previously advocated in the healthcare patient safety learning system literature. The standardisation-contextualisation tension constitutes a structural design challenge unique to the international level.

### Strengths and limitations

The cross-industry comparative design is a key strength: recruiting experts from civil aviation, maritime, and railway alongside healthcare generated perspectives not available from within healthcare alone and produced design insights grounded in operational experience of international systems that already function. Framework analysis, with its explicit link between data and study aims and its accommodation of both deductive and inductive coding, was well suited to the applied, pragmatic paradigm of this research. Independent coding of 20% of transcripts by an experienced qualitative researcher on the supervisory team reduced the risk of interpretive subjectivity.

The principal limitation is the small sample size (n = 11), consistent with published expert elicitation studies in this domain (Benn et al., 2009) but limiting representativeness. Perspectives from low-and middle-income country healthcare systems are absent. Data were collected in 2019 and the international patient safety governance landscape has continued to evolve since then, including through the WHO Global Patient Safety Action Plan 2021–2030, which post-dates data collection. Findings are therefore foundational rather than prescriptive. The inverted power dynamic of the interview process may have limited the depth of exploration on some topics; this was mitigated through a structured guide and reflexive practice throughout.

### Interpretation within the context of the wider literature

The finding that feedback mechanisms are the most consistently failing functional requirement of current national patient safety learning systems is consistent with the existing literature (Hewitt et al., 2017; Stavropoulou et al., 2015); this study extends it to the international level, where the structural distance between incident, analysis, and recommendation recipient makes the feedback gap greater. Blame culture, anonymity, and ease of reporting as design prerequisites likewise echo the national patient safety learning system literature (Health Quality Ontario, 2017; Mahajan, 2010), while the present study demonstrates that these barriers are qualitatively amplified rather than simply reproduced at the international level.

Two findings have no precedent in the national patient safety learning system literature. Legislative protection for reporters, a cornerstone of aviation’s safety reporting culture since the 1990s, has not previously been advocated as a structural prerequisite for a healthcare patient safety learning system. The standardisation-contextualisation tension was articulated across multiple participants as a design challenge with no established solution in the existing literature. The characterisation of barriers as contingent rather than inherent (Weaver et al., 2013) is also theoretically significant: feasibility is not foreclosed by blame culture or privacy legislation if the governance architecture is designed to address them, consistent with the WHO Global Patient Safety Action Plan’s emphasis on systemic governance as the foundation for international safety data sharing (WHO, 2021).

### Implications for policy, practice, and research

A single mandated international governance authority with cross-national representation is identified as the enabling condition for the system’s credibility and analytical quality, analogous to the International Civil Aviation Organization’s role in aviation. Legislative protection for reporters should be a policy priority concurrent with system design. A dual-database architecture, one for reported incidents and one for safety solutions and recommendations, would address a structural gap in current national systems. Rare and emerging events, never events, and near-misses with high harm potential constitute an empirically grounded starting framework for incident scope.

The qualitative evidence base established by this study provided the foundation for a subsequent modified Delphi consensus study with 21 international healthcare experts representing all six continents, which tested these design requirements against a broader expert panel and achieved consensus on 83% of evaluated statements (Qasem et al., 2026). Future research should examine the views of frontline healthcare professionals and patient representatives, who are absent from the expert sample reported here, and should address governance structures, funding models, and mechanisms for equitable participation from low-and middle-income countries.

## Conclusions

This study provides the first cross-industry qualitative account of design requirements for an international healthcare patient safety learning system. Rare and emerging event detection is the primary distinctive value of the international level; feedback, analytical capability, and actionable recommendations are the core functional requirements; anonymity, ease of reporting, and a single governance authority are the essential features; and legislative protection for reporters is a novel enabling condition with no equivalent in current healthcare patient safety learning system policy. These findings provide an empirical foundation for system design and policy development in alignment with the WHO Global Patient Safety Action Plan 2021–2030.

## Declarations

### Contributorship

Jaafer Qasem: Conceptualisation; Data curation; Formal analysis; Investigation; Methodology; Project administration; Visualisation; Writing – original draft; Writing – review and editing. Adrian Edwards: Conceptualisation; Methodology; Supervision; Writing – review and editing. Fiona Wood: Formal analysis; Methodology; Supervision; Writing – review and editing. Andrew Carson-Stevens: Conceptualisation; Methodology; Supervision; Writing – review and editing.

### Ethics approval

This study was approved by the Cardiff University School of Medicine Research Ethics Committee (SMREC Reference Number: 18/70). All participants provided written informed consent prior to interview.

## Funding

This work was supported by the Government of the State of Kuwait through a doctoral scholarship awarded to J. Qasem. The funder had no role in study design, data collection and analysis, decision to publish, or preparation of the manuscript.

## Conflict of interests

No known conflict of interests.

## Acknowledgements

The authors thank all participants who gave their time and expertise to contribute to this study.

## Data availability

Anonymised data are available on reasonable request from the corresponding author.

## Patient and public involvement

One participant was recruited specifically as a patient and public involvement representative with expertise in patient safety. No patient or public co-design of the study was undertaken; the participant contributed as a key informant in the same capacity as all other participants.

## Overlapping work

The interview findings reported in this manuscript informed a subsequent Delphi consensus study (Qasem et al., 2026). The two studies are independent in data and methods; interview participants were eligible for invitation to the Delphi panel, and some overlap in participation may exist.

## Notes

### Competing Interest Statement

The authors have declared no competing interest.

### Author Declarations

This study was approved by the Cardiff University School of Medicine Research Ethics Committee (SMREC Reference Number: 18/70).

### Summary of Updates

This revision corrects the number of experts invited to participate, from 73 to 21, with the response and completion figures updated accordingly (18 replied, 11 completed, a response-to-completion rate of 61%), consistent with the original doctoral thesis. The phrase "safety intelligence" in the abstract has also been revised to "safety data and learning" for clarity. No other changes have been made to the manuscript. All findings, themes, and conclusions remain unchanged.

