## Supplementary File 1 for "International patient safety learning systems: a qualitative key informant interview study with safety-critical industry experts" (from another version of this preprint: version 1, 2026-07-28)

**Extract of analytical matrix: charted interview data by participant and topic area**

International patient safety learning systems: a qualitative key informant interview study with safety-critical industry experts

*Note.* The matrix below presents an extract of the charted interview data produced during Stage 4 of the framework analysis (Ritchie & Spencer, 1994), illustrating four participants (P2, P3, P6, and P11) selected to represent the range of industries, perspectives, and data density across the dataset. Each cell contains illustrative verbatim quotations from one participant within one of the seven topic areas derived from the interview guide; these topic areas formed the initial thematic framework from which the seven reported themes were derived through Stage 5 mapping and interpretation. Quotation marks indicate verbatim extracts from interview transcripts. N/A indicates that the participant did not address this topic area substantively in their interview. Participant codes are randomised and non-specific to preserve anonymity. The full matrix is available from the corresponding author on reasonable request.

| | **Topic 1 Purpose of an international IRLS** | **Topic 2 Key features of an international IRLS** | **Topic 3 Key functions of an international IRLS** | **Topic 4 Transferability of learning between organisations/countries** | **Topic 5 Enabling factors for an international IRLS** | **Topic 6 Barriers to the setup of an international IRLS** | **Topic 7 Patient safety incidents relevant for an international IRLS** |
|---|---|---|---|---|---|---|---|
| P2 | N/A | "Well-maintained database in which the recommendations are openly available for anyone. Recommendations should have a national follow-up in the country in which the recommendation was issued. Other countries should have a process to keep itself up to date of new recommendations. National databases should be separate and only the relevant information should be transferred to the international database."<br>"Access should be open for everyone to read the | "I prefer anonymity. Anonymity lowers the barriers in reporting. This way all information is better handled, available and easier to share when there are not restricting confidentiality issues." | N/A | N/A | "Main priority should be to develop simple enough system and get as many possible countries to be committed. Important question is, which organisation can be responsible for the system? Maybe within the EU or the UN. Someone should take strong leadership and invite organisations from each country to collaborate." | "In my opinion, at the international level there should be learning at three categories: all most serious incidents resulting in the death of several persons, clear technical problems that can be solved worldwide, and learning from statistics resulting in serious consequences." |

| | | | | | | | |
|---|---|---|---|---|---|---|---|
| | | information that helps organisations and individuals learn. Maybe there should be only one named responsible organisation in each country.” | | | | | |
| P3 | “I suppose, big picture-wise, it's learning from other people's mistakes before you make them yourself. It is that sharing of information, and it's an awareness of what else is going on.” | “A responsible organisation should be found in each nation — the commitment of participants, national processes to investigate and collect information, guidelines and definitions about what kind of information is wanted into the database.” | “I think they should be anonymous, or I think people should have the option to make them anonymous. I think the names, and the location, apart from maybe the country where it happened, isn't particularly relevant.” | “So you know, having an international conference or, getting the people around the table, they all feel that their voice is being heard — every word would be debated, and that's why you've got such a great document, that the wording is accepted in 190 states around the world.” | “I think everything should start with a good quality investigation.” | “I think the more transparent you can make that, the better.” | “Well, I don't think it needs to have resulted in harm, but it just has to have the potential for it.” |
| P6 | “For the international level, the main purpose is I think to detect rare and emerging incidents — the purpose of the system is to be less focused on the really big problems that we know exist, but more on rare or emerging incidents.” | N/A | “It's really important in that mechanism — you need clinicians, subject matter experts and human factor experts to interrogate and understand the data.”<br>“An agreed high-level classification system, not too detailed — it needs to be useable so that it doesn't take too long to make a report.” | N/A | “Probably the other thing that's missing there is the governance of it. So, you have to have overarching governance around that database and you need representation from the countries who are submitting data.” | “And it needs to be trusted, you know, from that community — and that governance process needs to have good representation from people.” | “For the international level, the main purpose is I think to detect rare and emerging incidents — it's rare or emerging incidents that are the primary focus.” |
| P11 | “I would say it is to get an overview of the most important risk areas and be able to learn across countries — it's not necessary to go into all kinds of incidents, but it's more of an overview to learn across countries.” | “The main priority is to get reliable data into the system and try to feed back to the policy makers, which I think is the most important perhaps for international — and then have different forms of feedback, and give priority to feedback.” | “Mandatory for some people to actually report.” | “To be able to translate knowledge into practice you need to take into account the contextual conditions in each country, so a lot of work will probably have to go into adapting learning information to be relevant for different countries.” | “There needs to be some kind of a common definition of incidents across countries.”<br>“Resources and competence to analyse the information.” | “It's an enabler depending on if you succeed or not.”<br>“Under-reporting, which is a barrier all over the place.” | N/A |
