## Supplementary File 2 for "International patient safety learning systems: a qualitative key informant interview study with safety-critical industry experts" (from another version of this preprint: version 1, 2026-07-28)

**Semi-structured interview guides (two versions)**

International patient safety learning systems: a qualitative key informant interview study with safety-critical industry experts

**Note on guide development and use:** Two structurally identical versions of the interview guide were developed: one for participants from healthcare backgrounds (Version A) and one for participants from other safety-critical industries — primarily civil aviation, maritime, and railway — (Version B). The versions cover the same seven topic areas but differ in ordering: Version A begins with the purpose of an international patient safety learning system before moving to features and functions, reflecting the healthcare expert's familiarity with the healthcare reporting context. Version B begins with essential features and functions, drawing first on the participant's industry-specific knowledge before moving to cross-industry and healthcare-specific topics. Both versions were developed iteratively from findings of a systematic literature review conducted as the first phase of the broader research programme and reviewed with the supervisory team. The guide was piloted with a patient safety academic prior to use and amended accordingly. Interviewer transition phrases and contextual prompts are included to reflect the semi-structured approach. Bracketed text indicates in-the-moment adaptations to participant responses.

### VERSION A: Healthcare expert

#### Opening

Hi [confirm identity]. Thank you for making some time to allow me to interview you.

Before we start, would you mind confirming how much time you have?

I have reviewed research articles and relevant documents, but I recognise that not everything gets into the academic literature, and you, as an expert, have a unique perspective.

As mentioned in the shared participant information sheet, I am conducting this interview as part of a research project developing and testing an international incident reporting and learning system for healthcare.

There are six main topics that emerged from the reviewed research articles that we will discuss. Your input will help in refining the results of the review and in making changes accordingly.

*Note to interviewer: Confirm verbal consent if written consent form has not yet been received; request return of form afterwards.*

#### Body

Let us explore the purpose of an international incident reporting and learning system…

#### Topic 1: Purpose of an incident reporting and learning system

| |
|---|
| In your opinion, what do you consider the purpose of an incident reporting and learning system at an international level? |
| Please could you provide an example of such a system, or a system that you think might be closely related? |

Moving on…

**Topic 2: Components of an incident reporting and learning system**

| |
|---|
| In your opinion, what are the main components of an international incident reporting and learning system? |
| Let us examine each of those components in turn. Please could you describe [term used by the participant]? |

**Topic 3: Essential features of an incident reporting and learning system**

| |
|---|
| What do you believe is relevant feedback to be shared at the international level? |
| Next, would you mind sharing some relevant examples of such feedback? |
| Moving on, what features do you feel are essential for an international incident reporting and learning system? |
| [Follow-up if database is mentioned]: That is interesting — now, what should the database include? [e.g. reported incidents, safety solutions, or both?] |
| What should be the main priorities of an international incident reporting and learning system? |

Moving on…

**Topic 4: Enabling factors for the transfer of learning**

| |
|---|
| In your opinion, what do you believe is necessary to enable the transfer of learning at an international level? |

**Topic 5: Transferability**

| |
|---|
| In your opinion, is there an incident reporting and learning system that can be transferred and applied to healthcare at the international level? |
| Please describe it briefly. |

Moving on to the last topic…

**Topic 6: Barriers and enablers to setting up an international incident reporting and learning system**

| |
|---|
| In your opinion, what are the main enabling factors for setting up an incident reporting and learning system at the international level? |
| Let us examine each of those factors in turn. Please could you describe [term used by the participant]? |
| Next, in your opinion, what are the main barriers for setting up an incident reporting and learning system at the international level? |
| Let us examine each of those barriers in turn. Please could you describe [term used by the participant]? |

Moving on to the final topic…

**Topic 7: Patient safety incidents relevant for an international incident reporting and learning system**

| |
|---|
| In your opinion, what safety incidents are deemed essential for learning at the international level? |

**Closing**

Thank the participant for their time.

I have learned a great deal from this conversation. Are you happy for me to acknowledge you in any publication arising from this work? Please be assured that no quotations will be directly attributable to you. Goodbye.

**VERSION B: Non-healthcare safety-critical industry expert (civil aviation, maritime, railway)**

**Opening**

Hi [confirm identity]. Thank you for making some time to allow me to interview you.

Before we start, would you mind confirming how much time you have?

I have reviewed research articles and relevant documents, but I recognise that not everything gets into the academic literature, and you, as an expert, have a unique perspective.

As mentioned in the shared participant information sheet, I am conducting this interview as part of a research project developing and testing an international incident reporting and learning system for healthcare.

There are six main topics that emerged from the reviewed research articles that we will discuss. Your input will help in refining the results of the review and in making changes accordingly.

*Note to interviewer: Confirm verbal consent if written consent form has not yet been received; request return of form afterwards.*

**Body**

I have read Annexes 13 and 19, as well as the Safety Management Manual, and would like to know your opinion about the following…

*Note to interviewer: This version opens with features and functions, drawing first on the participant's industry expertise before moving to cross-industry and healthcare-specific topics.*

**Topic 1: Essential features of an incident reporting and learning system**

| |
|---|
| What do you believe is relevant feedback to be shared at the international level? |
| Next, would you mind sharing some relevant examples of such feedback? |
| Moving on, what features do you feel are essential for an international incident reporting and learning system? |
| [Follow-up if database is mentioned]: That is interesting — now, what should the database include? [e.g. reported incidents, safety solutions, or both?] |
| What should be the main priorities of an international incident reporting and learning system? |

Moving on, let us explore the purpose of an international incident reporting and learning system…

**Topic 2: Purpose of an incident reporting and learning system**

| |
|---|
| In your opinion, what do you consider the purpose of an international incident reporting and learning system? |
| Please could you provide an example of such a system, or a system that you think might be closely related? |

**Topic 3: Components of an incident reporting and learning system**

| |
|---|
| In your opinion, what are the main components of an international incident reporting and learning system? |
| Let us examine each of those components in turn. Please could you describe [term used by the participant]? |
| When you think about safety, what does 'learning' mean to you? |

From the research, there was no clear agreement on whether reporting safety incidents should be confidential or anonymous, particularly at the international level…

| |
|---|
| When it comes to reporting safety incidents at an international level, which is more relevant: confidentiality or anonymity? |
| Could you explain the reasoning behind your preference? |

Next, regarding access to an international incident reporting and learning system…

| |
|---|
| Who do you believe should have access to the system? |
| And how would [identified person] be enabled to access the system? |

**Topic 4: Enabling factors for the transfer of learning**

| |
|---|
| In your opinion, what do you believe is necessary to enable the transfer of learning at an international level? |

**Topic 5: Transferability**

| |
|---|
| In your opinion, is there an incident reporting and learning system that can be transferred and applied to healthcare at the international level? |
| Please describe it briefly. |

Moving on to the last topic…

**Topic 6: Barriers and enablers to setting up an international incident reporting and learning system**

| |
|---|
| In your opinion, what are the main enabling factors for setting up an incident reporting and learning system at the international level? |
| Let us examine each of those factors in turn. Please could you describe [term used by the participant]? |
| Next, in your opinion, what are the main barriers for setting up an incident reporting and learning system at the international level? |
| Let us examine each of those barriers in turn. Please could you describe [term used by the participant]? |

Moving on to the final topic…

**Topic 7: Patient safety incidents relevant for an international incident reporting and learning system**

| |
|---|
| In your opinion, what safety incidents are deemed essential for learning at the international level? |

**Closing**

Thank the participant for their time.

I have learned a great deal from this conversation. Are you happy for me to acknowledge you in any publication arising from this work? Please be assured that no quotations will be directly attributable to you. Goodbye.

*Note.* Both versions cover the same seven topic areas. Differences in ordering and initial framing reflect each participant group's primary professional context. Bracketed text represents in-the-moment adaptations to participant responses rather than fixed scripted questions, consistent with the semi-structured approach (Bearman, 2019; DeJonckheere & Vaughn, 2019).
